# Development and use analysis of ‘gestioemocional.cat’, a web app for promoting emotional self-care and access to professional mental health resources during the covid-19 pandemic

**DOI:** 10.1101/2021.08.11.21261793

**Authors:** SG Fidel-Kinori, G Carot-Sans, A Cuartero-Barbanoj, D Valero-Bover, J Piera-Jimenez, R Romà-Monfà, E Garcia-Ribatallada, P Perez-Sust, J Blanch-Andreu, JA Ramos-Quiroga

## Abstract

**Background:** Quarantines and nationwide lockdowns dictated for containing the spread of the COVID-19 pandemic may lead to distress and increase the frequency of anxiety and depression symptoms among the general population. During the national lockdown of the first wave of the COVID-19 outbreak in Spain, we developed and launched a Web App (Gestioemocional.cat) to promote emotional self-care in the general population and facilitate contact with healthcare professionals.

**Methods:** Gestioemocional.cat targeted all individuals aged 18 years or more and was designed by adapting the contents of a mobile App for adjuvant treatment of post-traumatic stress disorder (i.e., the PTSD Coach App) to the general population and the pandemic/lockdown scenario. We retrospectively assessed the utilization pattern of the Web App using data systematically retrieved from Google Analytics. Data were grouped into three time periods, defined using a join point analysis of COVID-19 incidence in our area: first wave, between-wave period, and second wave.

**Results:** The resulting Web App, maintains the navigation structure of the PTSD Coach App, with three main modules: tools for emotional self-care, a self-assessment test, and professional resources for on-demand contact. The self-assessment test combines the Patient Health Questionnaire-2 (PHQ-2) and the 7-item Generalized Anxiety Disorder Scale (GAD-7) and offers professional contact in the advent of a high level of depression and anxiety; contact is prioritized according to a screening questionnaire administered at the time of obtaining individual consent to be contacted. The tools for emotional self-care can be accessed either on-demand or symptom-driven. The utilization analysis showed a high number of weekly accesses during the first wave. In this period, press releases regarding critical events of the pandemic progression and government decisions on containment measures were followed by a utilization peak, irrespective of the sense (i.e., positive or negative) of the information. Positive information pieces (e.g., relaxation of containment measures due to a reduction of COVID-19 cases) resulted in a sharp increase in utilization immediately after information release, followed by a successive decline in utilization. The second wave was characterized by a lower and less responsive utilization of the Web App.

**Conclusions:** mHealth tools may help the general population coping with stressful conditions associated with the pandemic scenario. Future studies shall investigate the effectiveness of these tools among the general population―including individuals without diagnosed mental illnesses―and strategies to reach as many people as possible.

## 1 Introduction

In the last months, most countries worldwide have experienced uncontrolled outbreaks of the coronavirus disease 2019 (COVID-19) soon after reporting the first case. The rapid spread of its causing agent, the severe acute respiratory syndrome coronavirus 2 (SARS-CoV-2), has led to an unprecedented overburdening of healthcare systems, prompting many governments to dictate strict lockdowns for containing the outbreak.

While such indiscriminate strategies have succeeded in containing SARS-CoV-2 spread and reducing mortality (Flaxman et al., 2020), evidence warns on many psychological harms potentially associated with quarantines (Mohamad Marzuki et al., 2019), particularly among older and dependent people and those with previous psychiatric pathologies (Arango, 2020a; Holmes et al., 2020; Yang et al., 2020). Alongside the quarantine-specific stressors, the uncertainty associated with the pandemic scenario and impossibility of accompanying loved ones during their hospital stay or end-of-life stage has contributed to psychological overwhelming in many cases (Arango, 2020b).

The use of mobile- and web-based technologies for health (mHealth and eHealth, respectively) has been shown to improve access to healthcare services under confinement situations. In the last years, information and communication technologies (ICT) (including mHealth and eHealth solutions) for people with mental health needs have been gradually incorporated into routine care, also in social groups not considered regular digital consumers, such as older people (Hennemann et al., 2018; Schueller et al., 2018; Torous and Powell, 2015). A successful example of ICT-based solutions for mental health is the mobile App for managing post-traumatic stress disorder (i.e., the PTSD Coach app), developed by the Veterans Affairs National Centre for PTSD and the Department of Defence National Centre for Telehealth & Technology (Kuhn et al., 2014). The PTSD Coach app has been translated to many languages―including Spanish, a translation led by our team (Kuhn et al., 2018)―, and it is currently used in our Psychiatry Department as an adjuvant to face-to-face therapy for PTSD.

The Catalan Health Service, which provides universal healthcare to a population of 7.7 million inhabitants in North-East Spain, has a long tradition of implementing ICT solutions for promoting integrated care and increasing the efficiency and quality of healthcare delivery. The overloading of the healthcare system experienced during the COVID-19 outbreak boosted the implementation of various digital health strategies to counteract the collapse of many health resources (Sust et al., 2020). To our knowledge, by the time of the national lockdown in our country, no mHealth solutions had been developed for supporting people with mental health needs in a quarantine or lockdown context. Thus, considering the limited access of people to the healthcare system and the potential of ICT-based solutions to bridge these gaps, we aimed to develop a Web App platform for supporting emotional management among the general population during the COVID-19 outbreak in our country. The existence of the PTSD Coach App provided us with an opportunity to develop such a Web App quickly and offer it to the local population timely.

We present herein the development, main characteristics, and launch of the Web App, named gestioemocional.cat (“emotionalmanagement” in Catalan), and a retrospective analysis of its use throughout alternate periods of lockdown and ease of social distancing measures.

## 2 Methods

### 2.1 Design Objectives

The Web App (named Gestioemocional.cat; emotional management in the local language) was developed in response to a requested by the Catalan Health System and Catalan Ministry of Health to provide Catalonia citizens (North-East Spain) with support for coping with the emotional struggle associated with the COVID-19 pandemic and the national lockdown. The Web App was designed to seek two main objectives: (1) to provide users with tools and guidance for emotional self-assessment and emotional care, and (2) to establish a communication channel to reach healthcare professionals in the advent of worsening of mental health symptoms.

Gestioemocional.cat targeted all individuals aged 18 years or more with the capacity to use a mobile App or Website. Spain has a high smartphone penetration rate (De-Sola et al., 2019), for which the Web App had the potential to reach a significant part of the population. Still, technical requirements of the Web App were minimized to facilitate access. Furthermore, to achieve high acceptability and on-demand use of the Web App (Kummer and Schulte, 2019), we decided to store the least information possible and not keep the user’s historical records. Finally, the Web App could be directly downloaded from the institutional website of the Catalan Health Service to build trust on the tool.

#### 2.1.1 Web App Development

The government request for developing the Web App was made on March 26, 2020. A panel of experts consisting of two clinical psychologists with expertise in psychological trauma and crisis intervention and two psychiatrists of the Catalan Health System coordinated and oversaw the Web App development and content generation. Considering the limited time for developing the Web App and the effectiveness of the previously developed PTSD Coach App, we used it as a starting point to translate its contents and adapt them to a lockdown/pandemic scenario when necessary.

The panel of experts reviewed all items of the PTSD Coach App and grouped them into three categories: (1) items that could be translated into local languages without changes in their content, (2) items that required adaptation due to cultural differences, need for broadening the clinical scope to cover anxiety and depression symptoms, or adequateness to the quarantine/pandemic scenario, and (3) items to be removed for definite inappropriateness according to the intended use of the App. Owing to the urgency of Web App development, items review was based on the clinical criteria and expertise of panel members. Of note, two of them had contributed to translating the PTSD Coach App to Spanish and had, therefore, in-depth knowledge of the rationale behind each item. The resulting contents were used to building the Web App using the corporate colours and design of the Catalan Health System. The authors of the PSTD Coach App kindly provided permission to adapt and translate their App.

#### 2.1.2 Time Frame and Launching Strategy of the Web App

The first case of COVID-19 in Spain was officially reported in late January 2020. The Spanish government halted crowded events on March 9, 2020 and dictated a state of alarm and a national lockdown on March 13, 2020. The lockdown started a progressive easing on May 10, 2020 and was definitely lifted on June 20, 2020.

The Web App gestioemocional.cat was launched by the Catalan Health Department on April 15, 2020. Information regarding the Web App was disseminated using TV and radio advertisements, which were released through the channels of the public corporation.

#### 2.1.3 Utilization Analysis and Statistics

Data for utilization analysis of the Web App were gathered from Google Analytics. The main objective of the utilization analysis was to describe the number of users throughout the analysed period. We used a script to systematize queries for retrieving information on daily accesses to the Web App. For the longitudinal analysis of the Web App use, the number of events was transformed into a logarithmic scale and plotted against a timeline covering the period from the Web App launch on April 15, 2020, to dataset closure on December 16, 2020. Key events regarding outbreak progression (i.e., relevant press releases on epidemiological information and government decisions) and the time window of each wave of the outbreak were added to the plot. The two waves of the COVID-19 outbreak were defined using a join point regression analysis of publicly available data on COVID-19 incidence in our area. A permutation test with 0.05 confidence and 4,499 permutations was used as described elsewhere (Kim et al., 2000). Differences in the distribution of test scores on each of the priority levels in the three investigated periods (i.e., first wave, second wave, and between-waves period) were assessed using a Pearson’s Chi-squared test and setting the significance threshold at a two-sided alpha value of 0.05. The rest of the analyses were descriptive, and no other hypothesis tests were conducted. All analyses and plots were performed on R package (R Core Team, 2017).

## 3 Results

### 3.1 Platform Components

The gestioemocional.cat Web App has the same navigation structure as the PTSD Coach app. Table 1 summarizes the changes introduced to the PTSD Coach items to adapt them to the intended use of the gestioemocional.cat Web App; the resulting list of items was translated to the local languages (i.e., Spanish and Catalan)an also to English, for potential no native population. Figure 1 outlines the navigation structure of the Web App. The main menu provides the user with access to three sections: (1) resources for emotional management, (2) tools for self-assessment, and (2) professional resources.

**Table 1.**
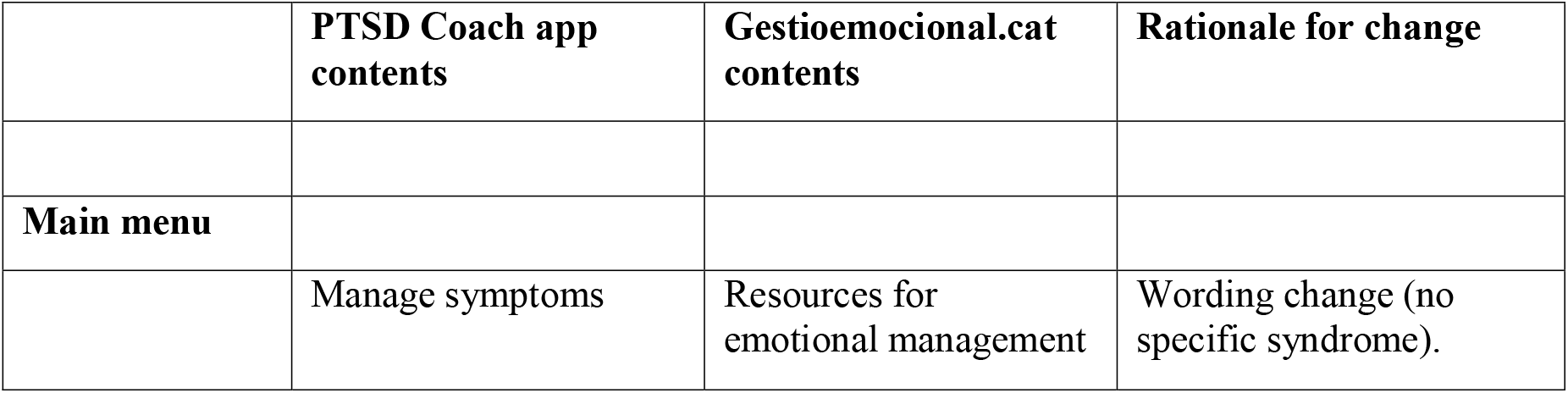
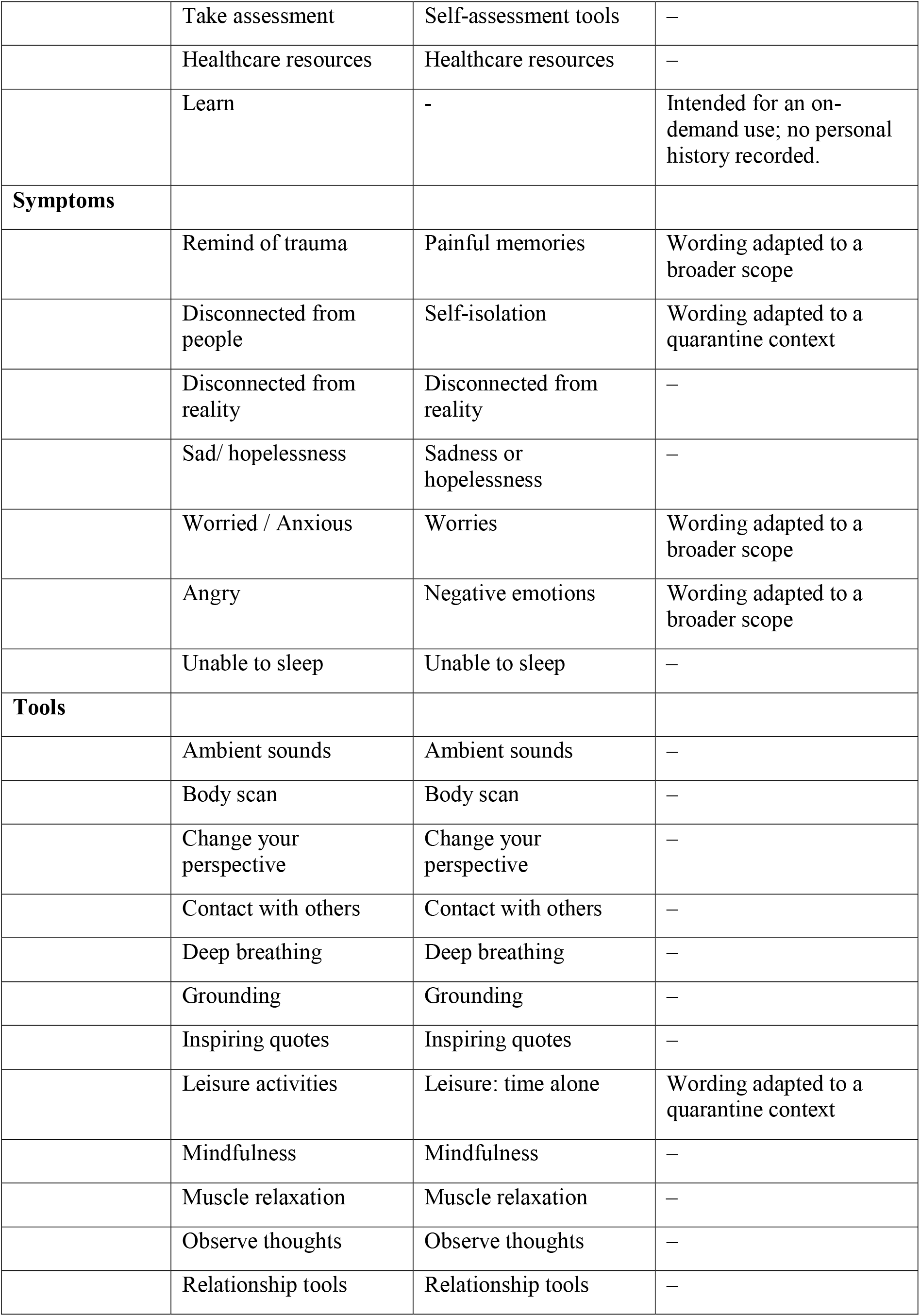
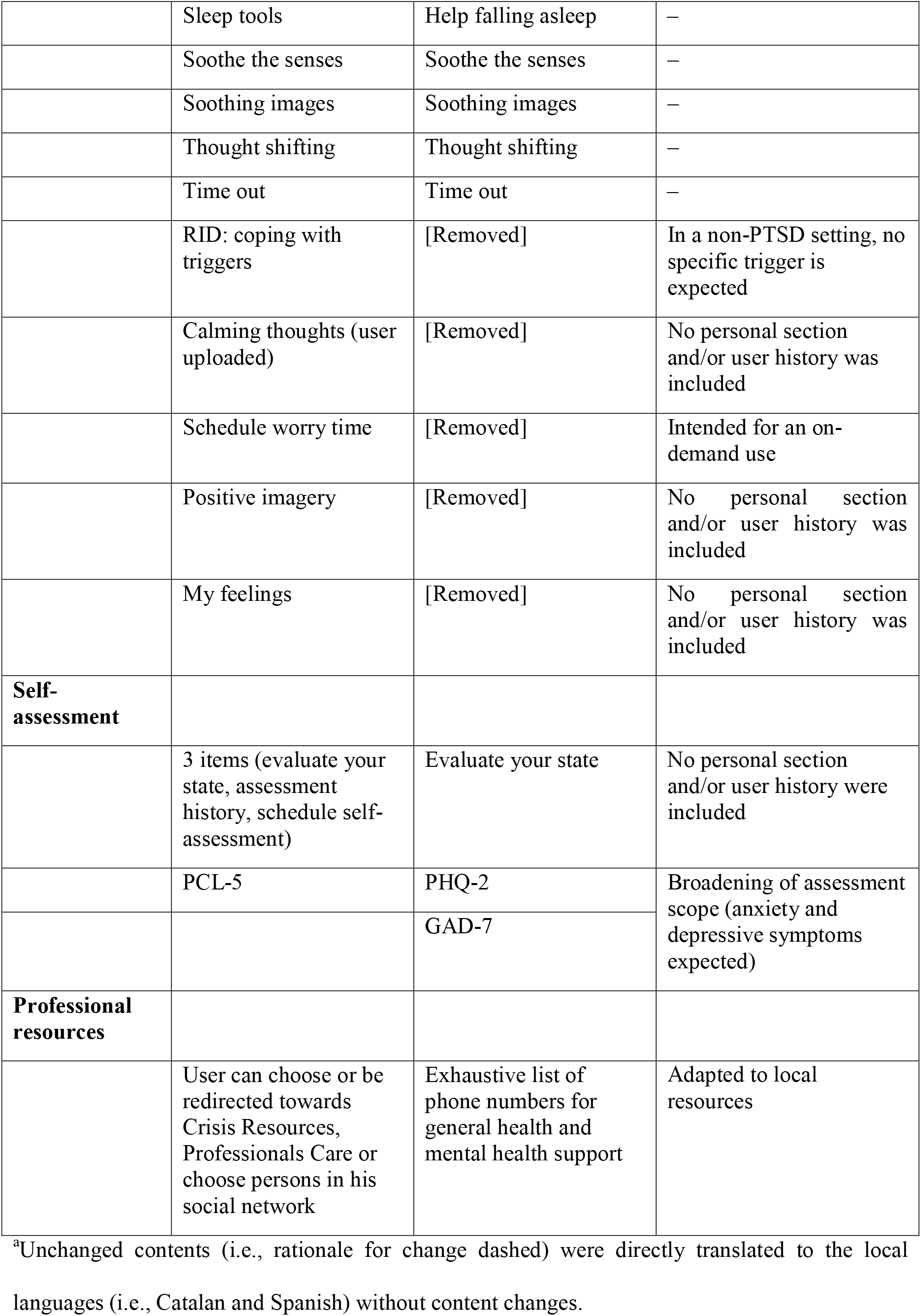
Summary of content changes for adaptation of the PTSD Coach app into gestioemociona.cat Web App.^a^

|  | **PTSD Coach app contents** | **Gestioemocional.cat contents** | **Rationale for change** |
| --- | --- | --- | --- |
| **Main menu** |  |  |  |
|  | Manage symptoms | Resources for emotional management | Wording change (no specific syndrome). |
|  | Take assessment | Self-assessment tools | – |
|  | Healthcare resources | Healthcare resources | – |
|  | Learn | - | Intended for an on-demand use; no personal history recorded. |
| **Symptoms** |  |  |  |
|  | Remind of trauma | Painful memories | Wording adapted to a broader scope |
|  | Disconnected from people | Self-isolation | Wording adapted to a quarantine context |
|  | Disconnected from reality | Disconnected from reality | – |
|  | Sad/ hopelessness | Sadness or hopelessness | – |
|  | Worried / Anxious | Worries | Wording adapted to a broader scope |
|  | Angry | Negative emotions | Wording adapted to a broader scope |
|  | Unable to sleep | Unable to sleep | – |
| **Tools** |  |  |  |
|  | Ambient sounds | Ambient sounds | – |
|  | Body scan | Body scan | – |
|  | Change your perspective | Change your perspective | – |
|  | Contact with others | Contact with others | – |
|  | Deep breathing | Deep breathing | – |
|  | Grounding | Grounding | – |
|  | Inspiring quotes | Inspiring quotes | – |
|  | Leisure activities | Leisure; time alone | Wording adapted to a quarantine context |
|  | Mindfulness | Mindfulness | – |
|  | Muscle relaxation | Muscle relaxation | – |
|  | Observe thoughts | Observe thoughts | – |
|  | Relationship tools | Relationship tools | – |
|  | Sleep tools | Help falling asleep | – |
|  | Soothe the senses | Soothe the senses | – |
|  | Soothing images | Soothing images | – |
|  | Thought shifting | Thought shifting | – |
|  | Time out | Time out | – |
|  | RID: coping with triggers | [Removed] | In a non-PTSD setting, no specific trigger is expected |
|  | Calming thoughts (user uploaded) | [Removed] | No personal section and/or user history was included |
|  | Schedule worry time | [Removed] | Intended for an on-demand use |
|  | Positive imagery | [Removed] | No personal section and/or user history was included |
|  | My feelings | [Removed] | No personal section and/or user history was included |
| **Self-assessment** |  |  |  |
|  | 3 items (evaluate your state, assessment history, schedule self-assessment) | Evaluate your state | No personal section and/or user history were included |
|  | PCL-5 | PHQ-2 | Broadening of assessment scope (anxiety and depressive symptoms expected) |
|  |  | GAD-7 |  |
| **Professional resources** |  |  |  |
|  | User can choose or be redirected towards Crisis Resources, Professionals Care or choose persons in his social network | Exhaustive list of phone numbers for general health and mental health support | Adapted to local resources |

<sup>a</sup>Unchanged contents (i.e., rationale for change dashed) were directly translated to the local languages (i.e., Catalan and Spanish) without content changes.
GAD-7: 7-item Generalized Anxiety Disorder Scale. PCL-5: PTSD Checklist for DSM-5. PHQ-2: patient health questionnaire-2. PTSD: post-traumatic stress disorder.

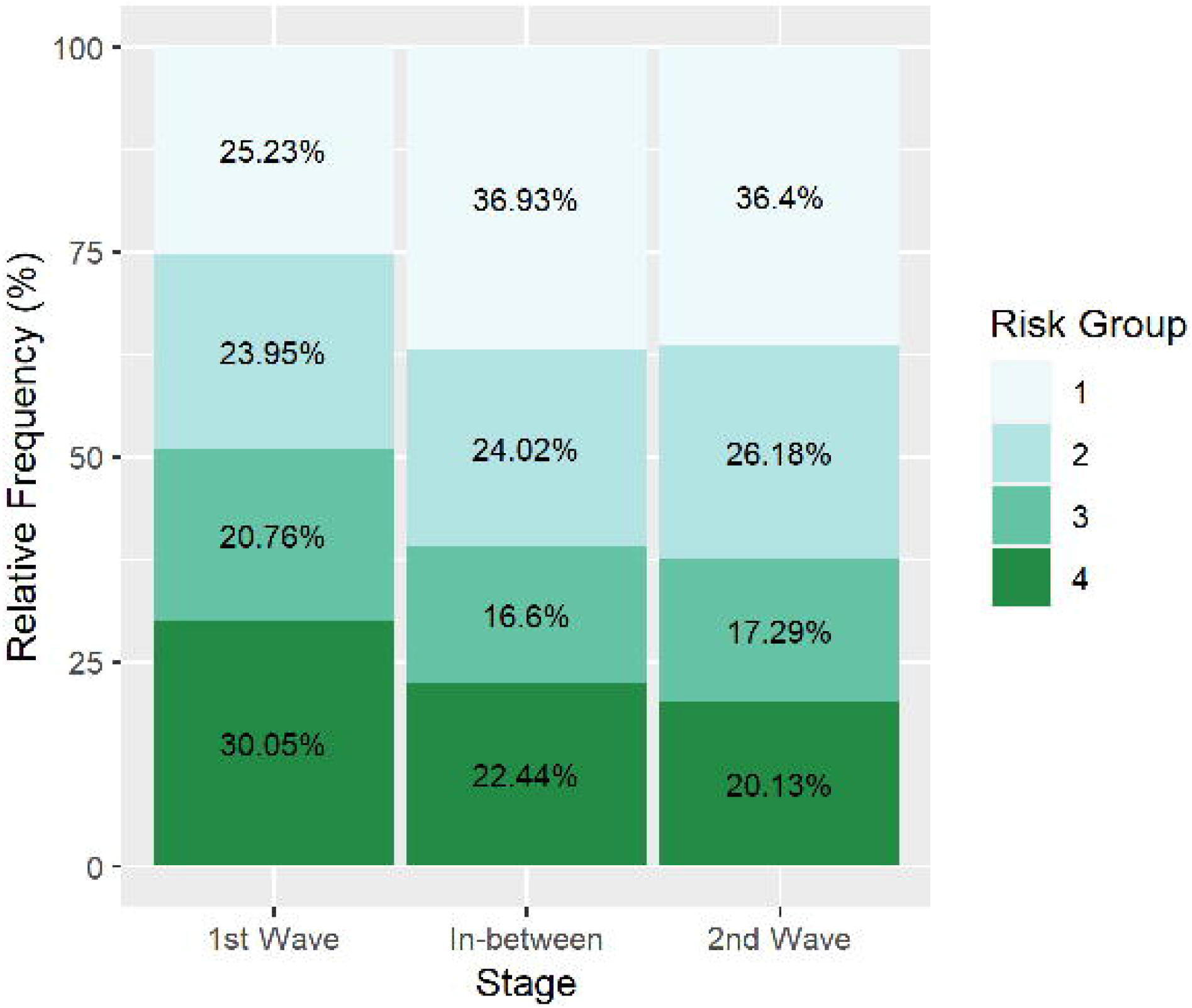

GAD-7: 7-item Generalized Anxiety Disorder Scale. PCL-5: PTSD Checklist for DSM-5. PHQ-2: patient health questionnaire-2. PTSD: post-traumatic stress disorder.

#### 3.1.1 Resources for emotional management

The section “resources for the emotional management” can be accessed through two pathways: symptom-guided and resource-type-guided (Figure 1). Symptoms include painful memories, self-isolation, disconnection from reality, sadness or hopelessness, worries, negative emotion, and sleeping disturbances. All these symptoms have been identified as very common during quarantines and lockdowns in the context of the COVID-19 pandemic (García-Álvarez et al., 2020; Xiong et al., 2020). Resource types include body scan (i.e., audio-guided), change your perspective (i.e., random text messages to rationalize distress thoughts), connect with others (i.e., list of ideas/tips for establishing interpersonal relationships under a quarantine context), deep breathing (i.e., audio-guided technique for emotional regulation trough breathing), grounding (i.e., list of tips for helping to be fully present for whatever is occurring right here and now), inspirational phrases, use of personal leisure time (i.e., list proposed activities to be performed during the lockdown), mindfulness exercises, progressive relaxation, observe thoughts (i.e., audio-guided techniques to help coping with distress thoughts), relationships improvements (i.e., list of tips to improve peer communication), help falling asleep (i.e., list of proposals for sleeping better), calm the senses, relaxing images, thought shifting (i.e., list of phrases to be repeated over five minutes using a countdown tool), and time-out (i.e., list of proposals to change the environment when remaining quarantined).

Irrespective of the pathway chosen to access the section “resources for the emotional management”, the Web App suggests a particular resource based on the user’s distress level, assessed using a single-item questionnaire (i.e., what is your distress level? [use the arrows to indicate how you feel]) and rated on a 0-10 scale. Users can select the distress level either stepwise―using up and down arrows―or using a distress thermometer with an intensity colour code (Figure 1)

#### 3.1.2 Self-assessment

The self-assessment section is based on two questionnaires for a mental health assessment with a Spanish version adapted and validated: the Patient Health Questionnaire-2 (PHQ-2) (Kroenke et al., 2003) and the 7-item Generalized Anxiety Disorder Scale (GAD-7) (Spitzer et al., 2006). The PHQ-2 questionnaire is intended as a first-step approach to identify depressive symptoms based on the frequency of depressed mood and anhedonia. While not recommended for depression diagnosis or monitoring, it is considered useful for screening for depression, which is the most common psychological conditions in clinical practice and research; individuals who screen positive should be further evaluated with the PHQ-9 to determine whether they meet the criteria for a depressive disorder (American Psychological Association, 2020). The Spanish version of the PHQ-2 questionnaire has shown good reliability and validity in identifying depressive symptoms in the primary care setting (Reuland et al., 2009). The GAD-7 questionnaire was developed for its use in primary care, and it has high sensitivity in identifying symptoms of the anxiety spectrum. The Spanish validation of the questionnaire has shown good reliability and validity (García-Campayo et al., 2010). To facilitate self-assessment, the two questionnaires were merged into a single questionnaire. The combination of the PHQ and GAD-7 questionnaires for a joint assessment of anxiety and depression has shown high internal reliability and strong convergent and construct validity when assessing its association with other mental health, quality of life, and disability measures (Kroenke et al., 2016). All items of the combined questionnaire refer to the frequency of symptoms within the past two weeks and are rated on a 4-point scale where 0 means “never”, 1 “less than seven days”, 2 “more than seven days”, and 3 “almost every day”.

The results of the two questionnaires are combined to provide four priority levels of care needs (Figure 2). Users with low scores in the PHQ-2 and GAD-7 scales (i.e., suggestive of mild symptoms of anxiety and depression) are encouraged to the routine use of self-management and self-assessment tools to either maintain (level 4) or improve (level 3) their emotional health status. Conversely, users with high scores in the PHQ-2 and GAD-7 scales (i.e., suggestive of severe anxiety and depressive symptoms) are offered phone contact with professionals of the emergency medical service. Users who agree to be contacted by a healthcare professional are redirected to a consent form, which collects minimum personal data. Phone contacts are prioritized based on the combined result of the self-assessment questionnaire (i.e., 1 or 2) and a screening form asking about six extreme mental health conditions: suicide ideation, loss of a loved one due to COVID-19, family member admitted to hospital due to COVID-19, living with someone at high risk of severe illness or with COVID-19 symptoms, current psychiatric treatment, and previous psychiatric treatment. Affirmative responses to these questions gave the highest priority, irrespective of the self-assessment questionnaire result (Figure 2).

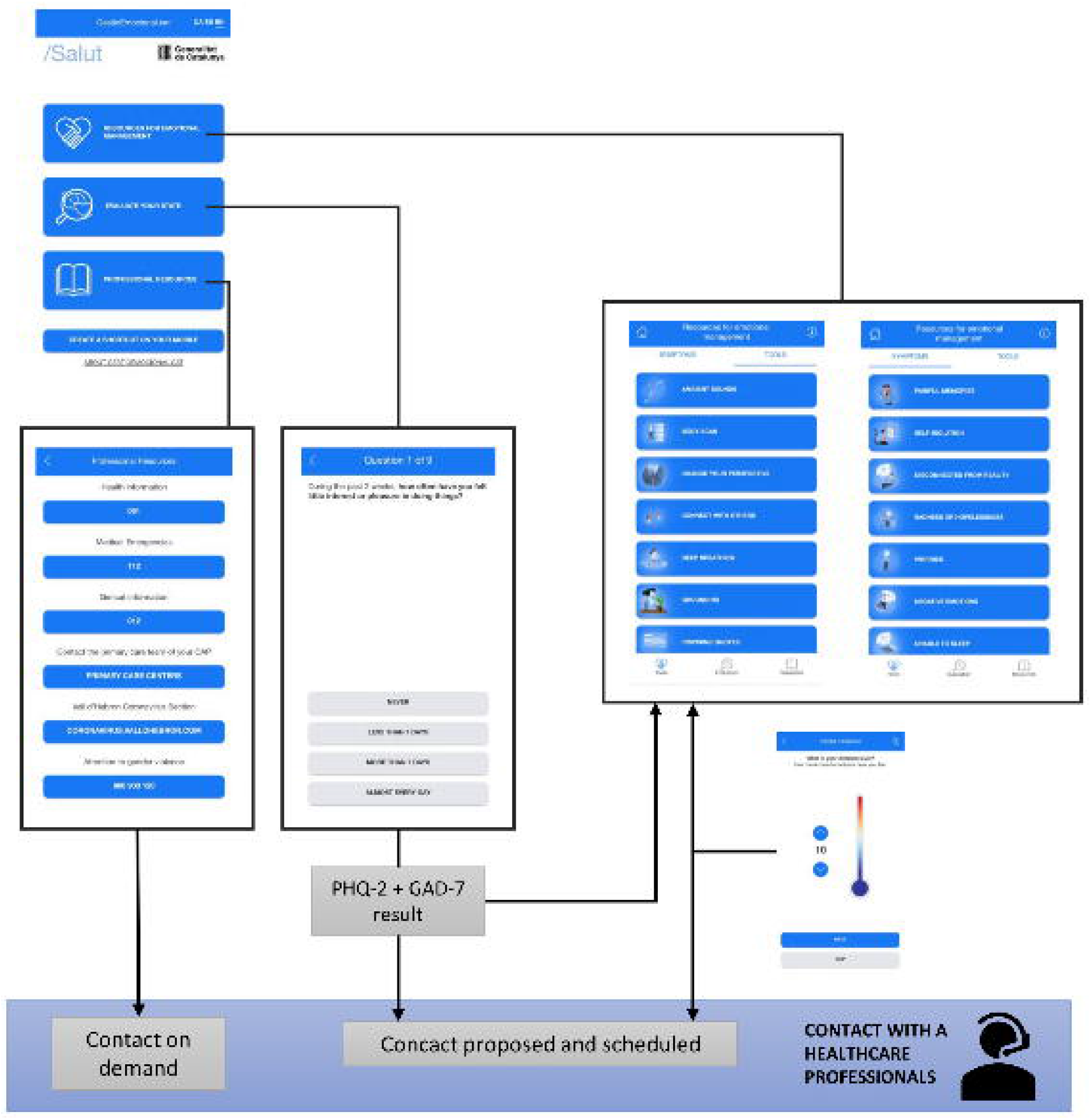

Scheduled phone calls are performed by the same professionals attending calls from people who reach the emergency medical service seeking mental health support. Once the phone contact has been established, healthcare professionals perform an independent assessment, irrespective of the contact source (i.e., direct call to emergency medical services or scheduled call through the Web App).

#### 3.1.3 Healthcare resources

The section “healthcare resources” allows the user to proactively contact community resources, including general health information, emergency medical service, general government information office, the user’s primary care centre, and the office for women under gender violence. Additional resources include the Galatea Foundation, aimed at providing support to healthcare professionals, the Official College of Psychologists, which can respond to population demands and transfer cases to public healthcare professionals, and the Vall d’Hebron Coronavirus Section, a web platform available 24 hours a day, 7 days a week for attending concerns related to emotional and social impact of the COVID-19 pandemic.

### 3.2 Utilization

Between April 15, 2020 (i.e., the launch date) and December 16, 2020, the Web App reported 582,826 accesses. Figure 3 shows the longitudinal analysis of the number of users and critical events of pandemic progress. During the last period of the national lockdown, the daily number of users peaked concomitantly with key news or government actions released by the press. Most of the press releases announcing changes on containment measures were immediately followed by a utilization peak, irrespective of the sense of these changes (i.e., increasing or easing restrictions). Immediately after announcing ease of restrictions (e.g., the start of the de-escalation plan), the daily number of users remarkably decreased. The decrease was particularly abrupt when Barcelona, the most populated area, entered the first phase of the de-escalation plan.

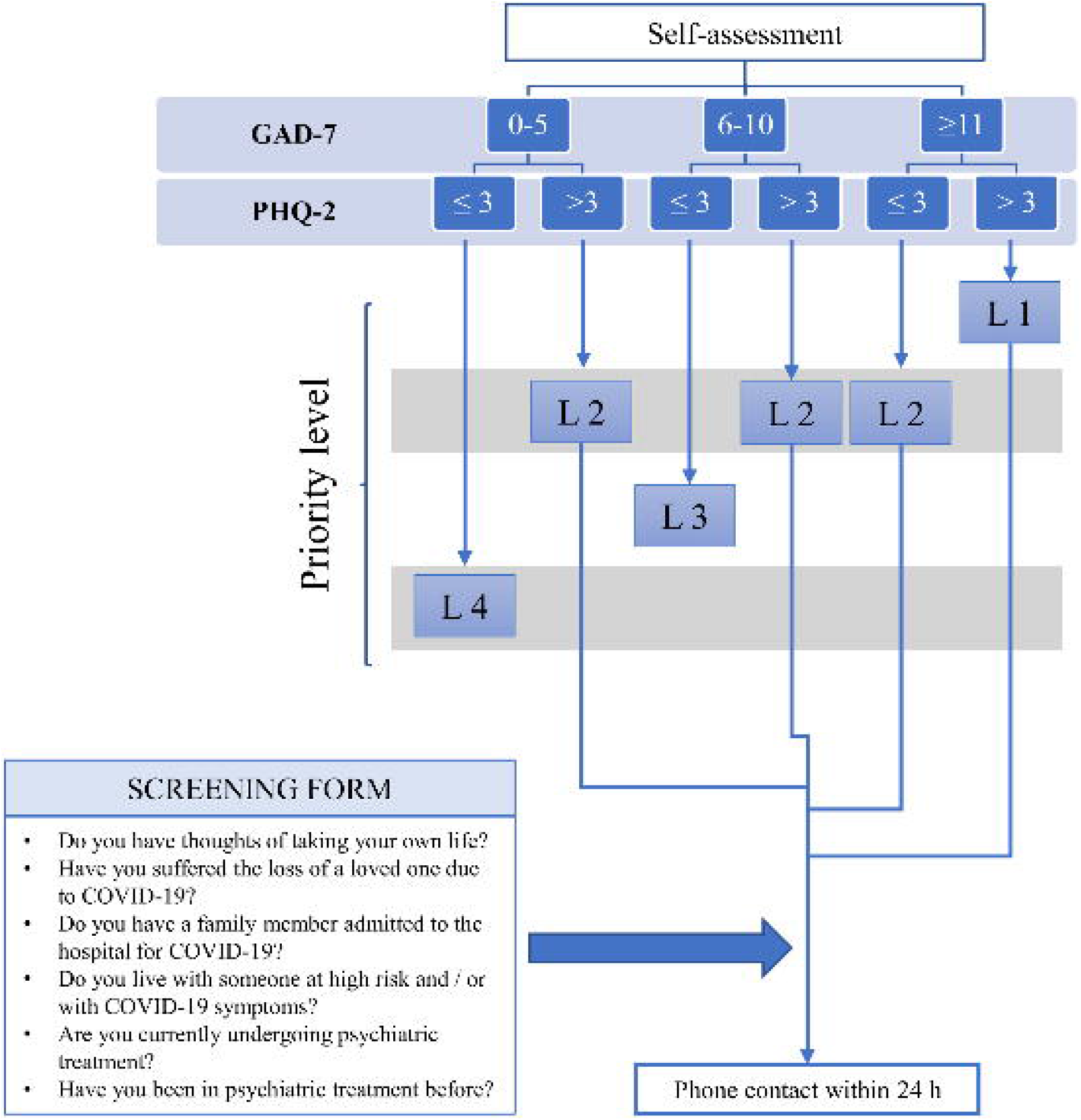

The number of users during the second wave of the outbreak was persistently lower than that in the first wave. During this period, Web App utilization was less sensitive to the announcement of government measures.

### 3.3 Progression of self-assessment

During the investigated period, the Web App recorded 142,337 self-assessment tests: 124,087 (87.2%) during the first wave, 8,510 (6.0%) during the period between waves, and 9,740 (6.8%) during the second wave. The Chi-squared test revealed significant differences regarding the distribution of percentages across the priority levels (obtained from the self-assessment module) between the investigated periods (Figure 4). The percentage of tests scoring for priority levels 1 or 2 (associated with more severe symptoms of anxiety and depression) was higher in the second wave and between-waves periods. Overall, the offer of professional help was accepted in 8,588 cases (11.4% of all cases with priority level 1 or 2 and, therefore, triggering the message of professional help offer).

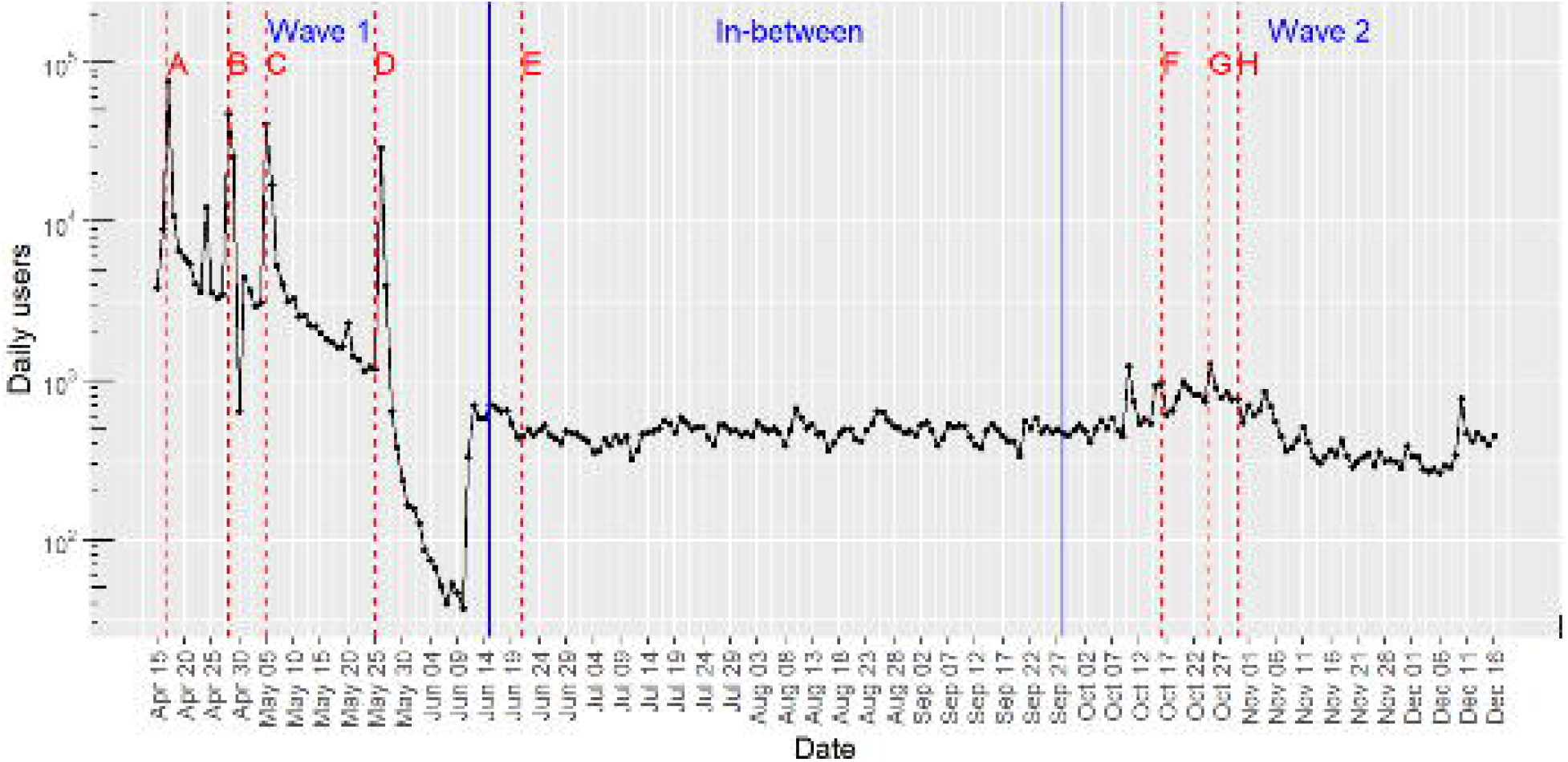

## 4 Discussion

In this manuscript, we describe a Web App aimed at providing the general population with self-care and self-assessment tools for emotional health during the lockdown associated with the COVID-19 pandemic and establishing a new communication channel between the population and healthcare professionals. In addition to reporting the characteristics of the Web App, we analysed the utilization pattern of the Web App during the first ten months of the COVID-19 outbreak in Spain. The frequency of use of the Web App was particularly high during the first wave of the outbreak in Spain and progressively declined until reaching a baseline activity below 1,000 weekly accesses after halting the national lockdown. During the first wave, the number of accesses peaked after press releases associated with the pandemic progression or changes in the containment measures dictated by the government. Interestingly, positive news (e.g., relaxation of social distancing measures) were immediately followed by a sharp increase in utilization and a successive decline. The between-waves and second wave periods were characterized by a less responsive pattern of Web App users.

The self-assessment module of the Web App, designed to work as a sentinel for detecting severe mental health symptoms and proactively offer direct contact with healthcare professionals, was mostly used during the first wave. However, the scores of the self-assessment questionnaire were higher after the first wave, suggesting a higher proportion of people with mental healthcare in the second wave and between-waves periods.

To our knowledge, by the time of developing the Web App, no other mHealth solutions for promoting emotional self-care in the general population during a lockdown or quarantine situation had been launched or published. The closest example of a mHealth tool that matched our aim was the PTSD Coach App, which was innovative in the sense that users can access tools through two alternative pathways: on-demand and symptom-driven (Kuhn et al., 2014). Although the PTSD Coach App was designed as adjuvant to face-to-face therapy for PTSD, its authors suggested that it could also help people not receiving psychological treatment (Kuhn et al., 2017; Ruzek et al., n.d.). Further experiences also showed good usability and ease of cultural adaptation of the App (Kuhn et al., 2018), encouraging us to adapt its contents to a lockdown and pandemic scenario in our country.

Although we cannot directly compare the utilization patterns of our Web App with other mHealth or eHealth solutions for lockdown contexts, the trends observed in our analysis are consistent with information reported elsewhere. For instance, the increased consumption of alcohol and drug abuse reported during this period in our area (García-Álvarez et al., 2020) could likely be attributed to increased anxiety symptoms, grief processes, and difficulties associated with intensive cohabitation (Ben-Zur and Zeidner, 2009; Boscarino et al., 2006). Likewise, other countries have reported increased demand for attention from people with mental health needs during nationwide lockdowns associated with the COVID-19 pandemic (Liu et al., 2020; Wang et al., 2020).

The design of our retrospective analysis precludes drawing strong conclusions regarding the reasons behind the observed trend towards a less responsive utilization of the Web App in the second wave of the outbreak. However, this finding is consistent with the “pandemic fatigue” phenomenon highlighted by the WHO and defined as an expected and natural response to a prolonged public health crisis (World Health Organization (WHO), 2021). This type of reaction might correspond to M. Seligman’s Paradigm on learned helplessness, which describes a passive behaviour exhibited by a subject after enduring repeated aversive stimuli beyond their control. It is worth noticing, however, that self-assessment tests more often scored priority levels 1 or 2 during the between-wave and second wave periods, suggesting that people with higher mental health needs were more likely to retain the Web App utilization after the first wave.

A second objective of the Web App was to establish a new way of access to specialized healthcare professionals of the emergency department of the public healthcare system. The fact that 8,588 self-assessment tests ended up with a scheduled phone call with healthcare professionals for mental health support indicates that the Web App has emerged as an additional communication channel for seeking professional help. The number of phone contacts represents a low percentage (11.4%) of the total situations in which professional support was offered. However, this finding is consistent with current evidence, which suggests that most people with common mental health problems do not seek professional help (Bayer and Peay, 1997; Oliver et al., 2005).The development and launching of the Web App were strongly influenced by the urgency and overloading of the healthcare system, which challenged the design of an adequate assessment strategy of the platform. We chose the PTSD coach app because of the symptom overlapping between post-traumatic stress disorder and those reported elsewhere for lockdown situations, particularly regarding anxiety and depression. One of the advantages of the PTSD coach app was the body of evidence on its effectiveness for self-management of mental health needs. However, owing to the shift in the target population (i.e., from people diagnosed with post-traumatic stress disorder to the general population), effectiveness of our Web App cannot be assumed and should be confirmed in future studies. Of note, the complicated scenario in which the Web App should be assessed (i.e., quarantine context) will strongly challenge the trial conduct.

Our analysis was limited by the few data recorded and stored from the Web App, which precluded more exhaustive assessments of behaviour patterns among users (e.g., to discriminate between entries of new users and regular access of the same user). However, because of the negative correlation that often exists between App use/demand and permissions for accessing private data (Kummer and Schulte, 2019), we were afraid that collecting too much data from users might be a barrier to Web App utilization. Of note, the government announcement of launching a COVID-19 tracer app raised many privacy concerns among the population and was strongly criticized. In light of this general perception, we prioritized the acceptability of the Web App over the capacity of collecting personal information. As a result, our database for the retrospective analysis lacked important information that might have helped to understand behaviour patterns among users. One of the most remarkable consequences of this limitation is the lack of data at a user level, which precludes discriminating between entries of new users and regular access of the same user. More importantly, the lack of historical records of users prevents us from analyzing improvement or worsening of mental health state. Finally, we could not analyse Web App utilization according to socioeconomic status, which has been highlighted as a source of inequality in other aspects associated with access to COVID-19 resources (Amengual-Moreno et al., 2020; Baena-Díez et al., 2020). Nevertheless, while all these limitations compromise analytical approaches of the Web App, the absence of personal questions and no need for signing in are likely to promote the platform use, in line with the overarching goal of the project, which was to help the population during the stressful circumstances of the lockdown.

## 5 Conclusions

In the first year of the COVID-19 pandemic, various authors have stressed the need to promote original and creative ways to help people cope with the new and stressful conditions behind the pandemic (Inkster et al., 2020; Torous et al., 2020). We did not want to miss the opportunity to adapt mHealth tools with proven efficacy in self-management of stressful situations to the pandemic scenario. Future studies shall investigate to what extent this tool helps people coping with the pandemic scenario and address strategies for making as many people as possible aware of this resource.

## Data Availability

The datasets generated during this study are available from the corresponding author on reasonable request.

## Abbreviations

App: Application
COVID-19: Coronavirus disease 2019
GAD-7: Generalized Anxiety Disorder Scale
ICT: Information and communication technologies
PHQ-2: Patient Health Questionnaire-2
PTSD: Post-traumatic stress disorder
SARS-CoV-2: Severe acute respiratory syndrome coronavirus 2
WHO: World Health Organization

## Acknowledgements

The authors would like to thank all the teams involved in the development of the technological solution and the delivery of the intervention from Servei Català de la Salut, Centre de Telecomunicacions i Tecnologies de la Informació, Sistema d’Emergències Mèdiques and Departament de Salut. We also thank the developers of the PTSD Coach app (particularly Drs. Jason E Owen and Erik Kuhn) for kindly permitting us to adapt the contents of their App.

